# Barriers and Facilitators to HIV Prevention Services Among Key Populations in Biakoye District, Ghana

**DOI:** 10.64898/2025.12.03.25341565

**Authors:** Eyram Kuma Hanu, Rita Ama Wurapa, Martin Tettey, Mavis Pearl Kwabla

## Abstract

Key populations such as sex workers, men who have sex with men (MSM), women who have sex with women (WSW), and people who inject drugs (PWID) bear a disproportionately high burden of HIV infection. Despite the availability of HIV prevention services in Ghana, access and utilization remain limited, particularly in rural settings. This cross-sectional study conducted in April 2025 assessed barriers and facilitators to HIV prevention service utilization among 43 key population members in Biakoye District. Data were collected through a structured questionnaire and analyzed using descriptive statistics, chi-square tests, and logistic regression. Participants had a mean age of 25.2 years; most were female (65.1%) and sex workers (72.1%). Although 93% had heard of HIV prevention methods, only 62.8% had ever used prevention services. Key barriers included lack of awareness (48.8%), fear of stigma or discrimination (48.8%), long waiting times (36.1%), transportation challenges (29.5%), and confidentiality concerns (34.9%). Higher education (aOR = 2.60, 95% CI: 0.41–4.73, p = 0.020) and knowledge of service locations (aOR = 1.55, 95% CI: 0.08–3.03, p = 0.039) were significantly associated with utilization. Findings highlight persistent structural and social barriers impeding service uptake among key populations and underscore the need for stigma reduction, improved accessibility, and community-based HIV prevention models in rural districts.

## INTRODUCTION

Human Immunodeficiency Virus (HIV) continues to be a major global health challenge, disproportionately affecting key populations (KPs) such as female sex workers (FSWs), men who have sex with men (MSM), women who have sex with women (WSW), and people who inject drugs (PWID). Globally, nearly half of individuals within KPs do not receive essential prevention services such as condoms, HIV testing, treatment linkage, harm reduction, or pre-exposure prophylaxis (PrEP) (1,2). In sub-Saharan Africa, KPs and their partners accounted for approximately 25% of new HIV infections in 2022 (3).

In Ghana, the national HIV prevalence is estimated at 1.7%, yet prevalence among KPs is substantially higher 11.1% among FSWs and 17.5% among MSM (4,5). Although national programs promote condom use, HIV testing, and PrEP, utilization is hindered by stigma, discrimination, confidentiality concerns, and limitations in rural health systems (6). Persistent gaps between awareness and consistent preventive behavior have also been observed, influenced by economic pressure, partner dynamics, and limited female-controlled prevention methods.

Barriers to HIV prevention among KPs are multidimensional, spanning individual, social, and structural determinants. Stigma, discrimination, inconvenient service hours, transportation difficulties, and limited-service availability have been widely reported (8,16). Conversely, facilitators such as community empowerment, peer-supported models, integrated service delivery, and trust in healthcare providers improve engagement with prevention services (16,23).

Rural districts such as Biakoye are understudied, and evidence on HIV prevention among KPs in these areas is limited. Understanding the knowledge, accessibility, utilization, barriers, and facilitators in this context is essential to inform targeted interventions. This study therefore assessed the barriers and facilitators to HIV prevention services among key populations in Biakoye District, Ghana.

## MATERIALS AND METHODS

### Study Design and Setting

A cross-sectional study was conducted from 1^st^ to 30^th^ April 2025 in Biakoye District, Oti Region of Ghana, a predominantly rural area with limited health infrastructure and decentralized HIV services.

### Study Population and Sampling

Participants were adults aged ≥18 years who self-identified as belonging to a KP group (FSWs, MSM, WSW, or PWID) and resided in the district. Snowball sampling through peer networks identified 43 participants.

### Data Collection

A structured interviewer-administered questionnaire captured socio-demographics, knowledge of HIV prevention, service accessibility, utilization patterns, and perceived barriers and facilitators.

### Data Analysis

Data were analyzed using Stata version 17. Descriptive statistics summarized participant characteristics. Chi-square tests examined associations between variables. Factors significant at p < 0.10 were entered into multivariable logistic regression to determine predictors of HIV prevention service utilization. Adjusted odds ratios (aORs) and 95% confidence intervals (CIs) were reported.

### Ethical Considerations

Ethical approval was obtained from the Ghana Health Service Ethics Review Committee (GHS-ERC: 022/03/25). Written informed consent was obtained, and confidentiality assured.

## RESULTS

### Socio-demographic Characteristics

Participants (N = 43) had a mean age of 25.2 years (SD = 4.9). Most were female (65.1%) and single (88.4%). Over half were aged ≥25 years (58.1%). More than a third had tertiary education (34.9%). Sex workers comprised the majority (72.1%). This is displayed in table 1 below

**Table 1:**
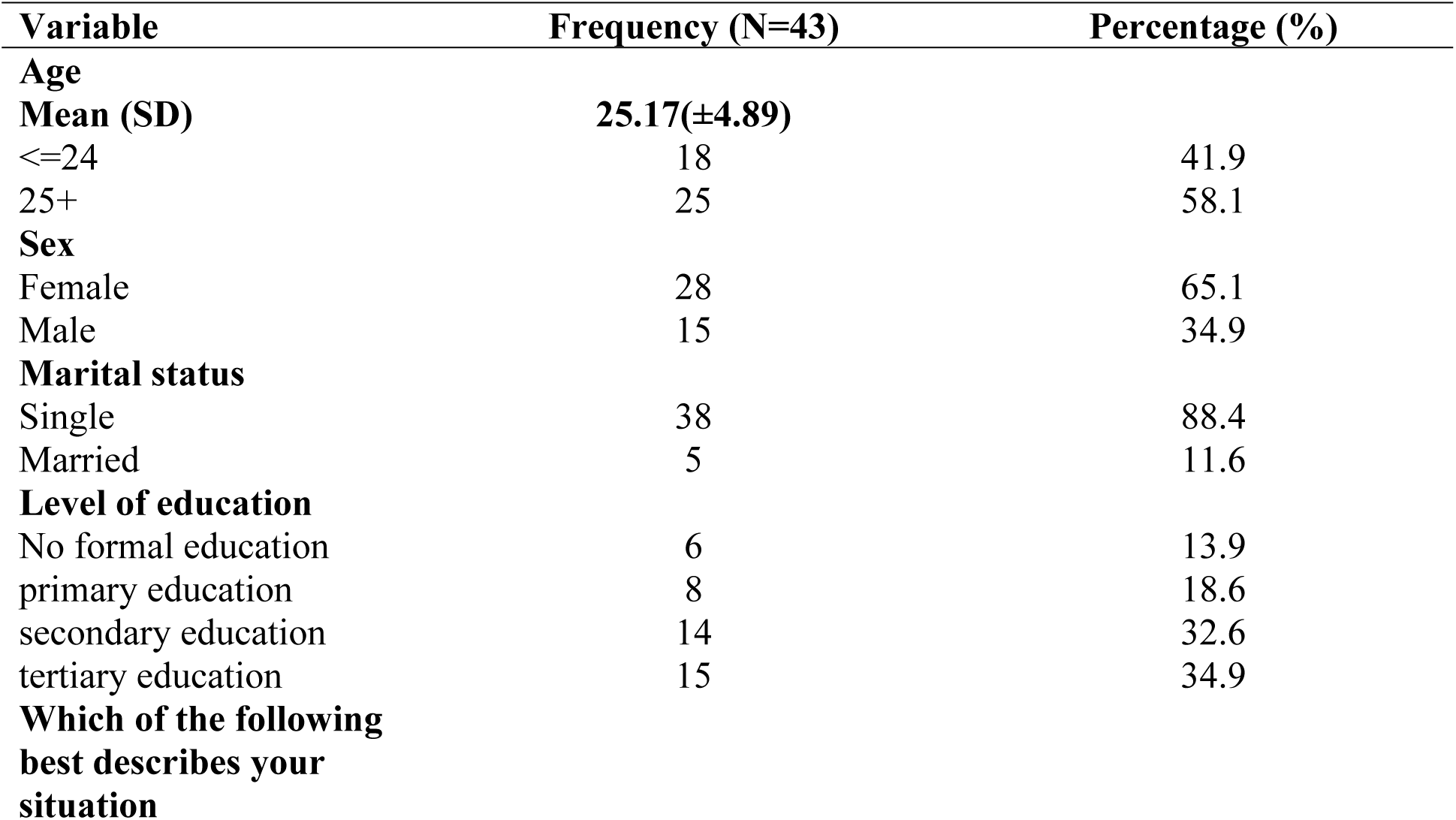
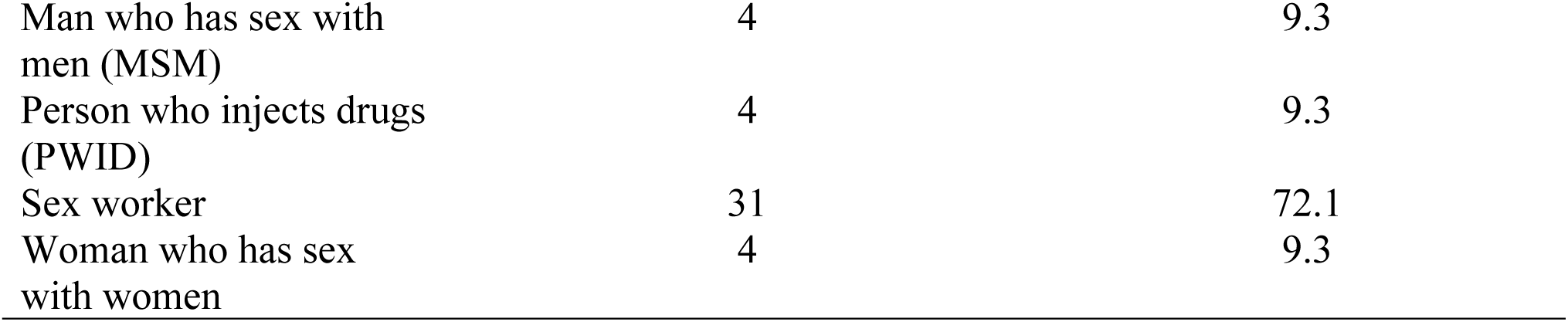
Socio-demographic Characteristics.

| Variable | Frequency (N=43) | Percentage (%) |
| --- | --- | --- |
| **Age** |  |  |
| Mean (SD) | 25.17(±4.89) | |
| $\leq 24$ | 18 | 41.9 |
| 25+ | 25 | 58.1 |
| **Sex** |  |  |
| Female | 28 | 65.1 |
| Male | 15 | 34.9 |
| **Marital status** |  |  |
| Single | 38 | 88.4 |
| Married | 5 | 11.6 |
| **Level of education** |  |  |
| No formal education | 6 | 13.9 |
| primary education | 8 | 18.6 |
| secondary education | 14 | 32.6 |
| tertiary education | 15 | 34.9 |
| **Which of the following best describes your situation** |  |  |
| Man who has sex with men (MSM) | 4 | 9.3 |
| Person who injects drugs (PWID) | 4 | 9.3 |
| Sex worker | 31 | 72.1 |
| Woman who has sex with women | 4 | 9.3 |

**Table 2:**
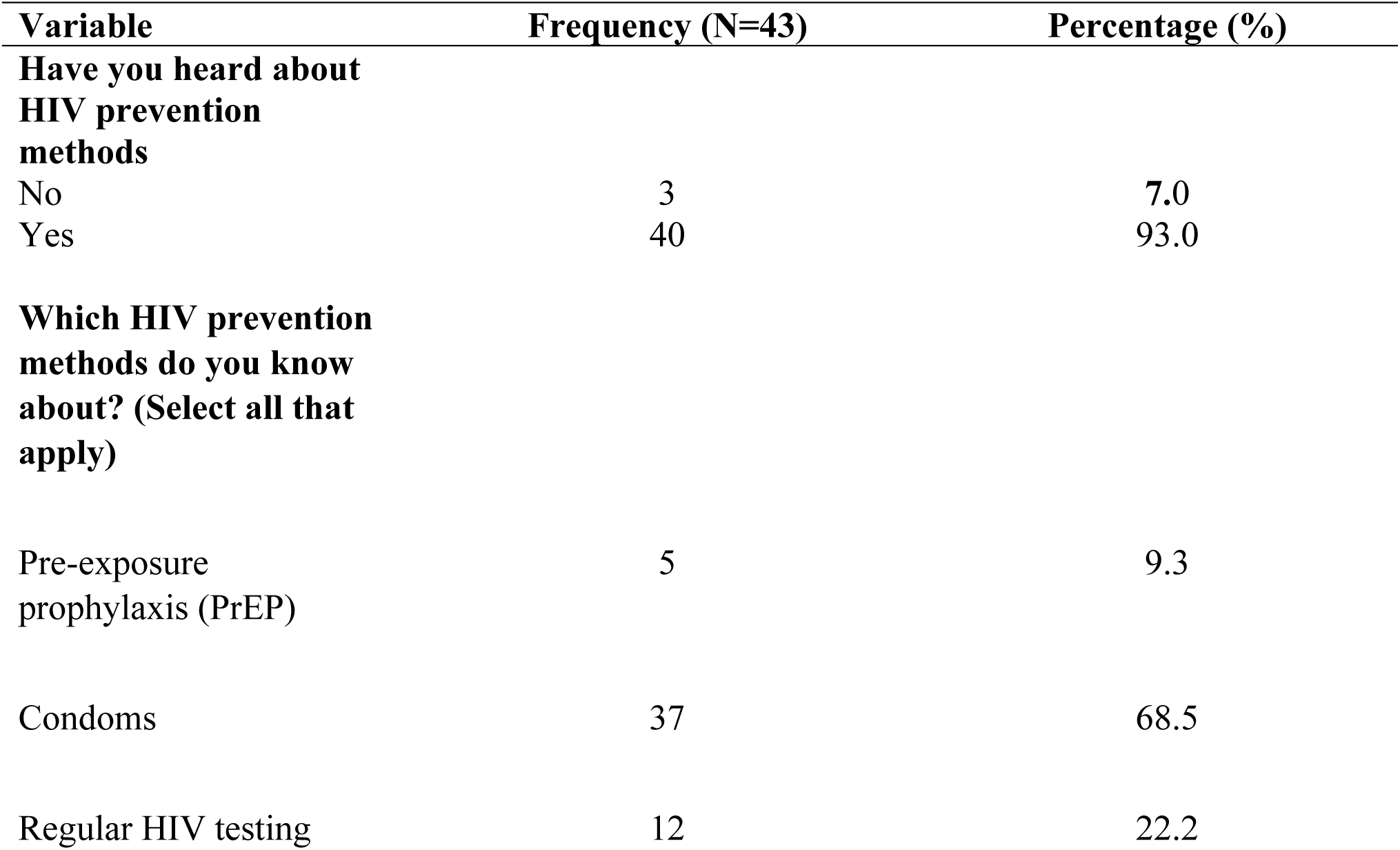
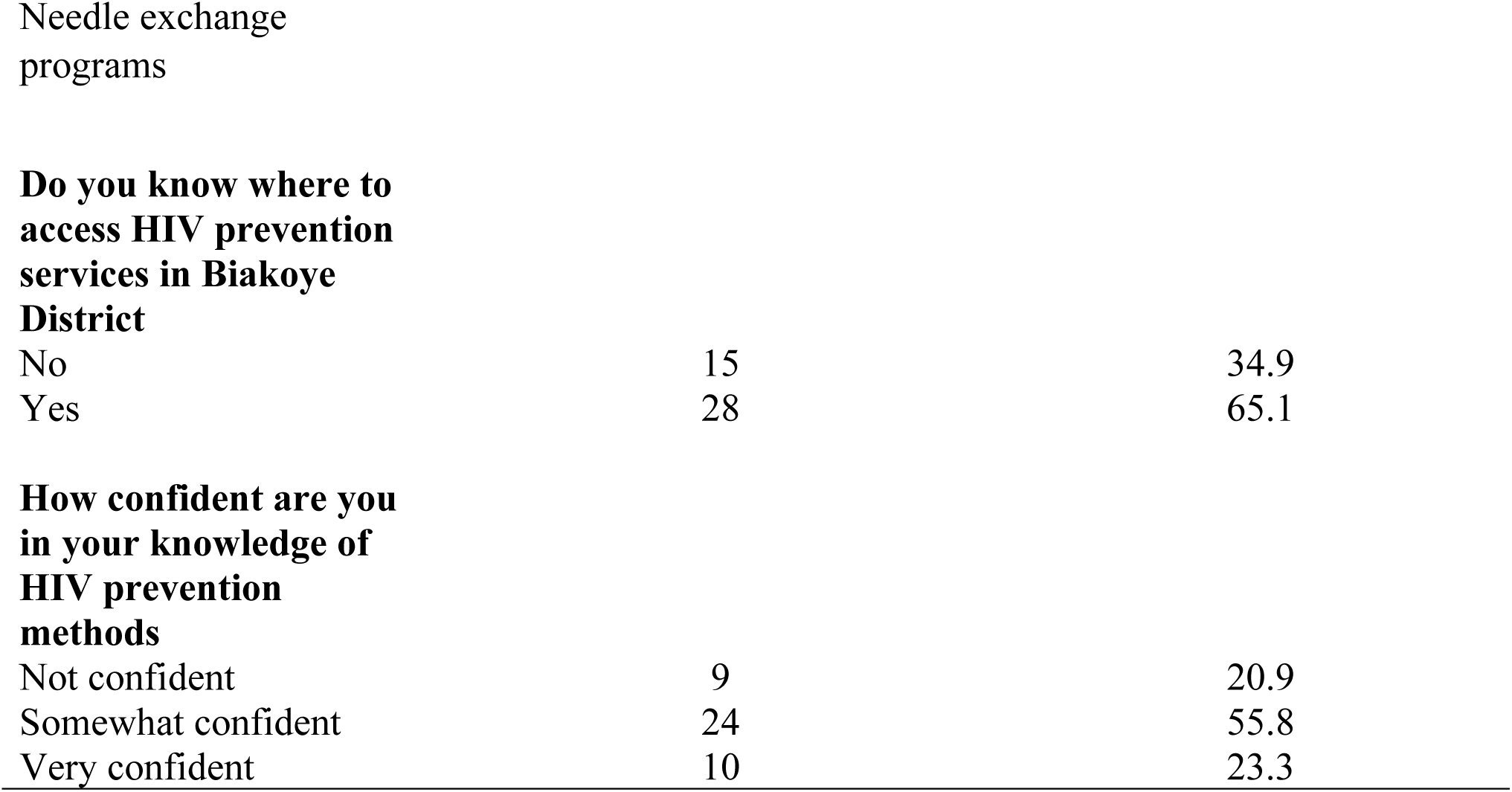
Knowledge of HIV Prevention.

| Variable | Frequency (N=43) | Percentage (%) |
| --- | --- | --- |
| **Have you heard about HIV prevention methods** |  |  |
| No | 3 | 7.0 |
| Yes | 40 | 93.0 |
| **Which HIV prevention methods do you know about? (Select all that apply)** |  |  |
| Pre-exposure prophylaxis (PrEP) | 5 | 9.3 |
| Condoms | 37 | 68.5 |
| Regular HIV testing | 12 | 22.2 |
| **Do you know where to access HIV prevention services in Biakoye District** |  |  |
| No | 15 | 34.9 |
| Yes | 28 | 65.1 |
| **How confident are you in your knowledge of HIV prevention methods** |  |  |
| Not confident | 9 | 20.9 |
| Somewhat confident | 24 | 55.8 |
| Very confident | 10 | 23.3 |

**Table 3:** Accessibility and Utilization of HIV Services.

| Variable | Frequency (N=43) | Percentage (%) |
| --- | --- | --- |
| **Have you ever used any**<br>**HIV prevention services** |  |  |
| No | 16 | 37.2 |
| Yes | 27 | 62.8 |
| **If yes, which services have**<br>**you used? (Select all that**<br>**apply)** |  |  |
| PrEP | 1 | 2.6 |
| Condoms | 27 | 71.1 |
| Regular HIV testing | 10 | 26.3 |
| **How frequently do you access these services** |  |  |
| Occasionally (24 times a year) | 9 | 33.3 |
| Rarely (once a year or less) | 6 | 22.5 |
| Regularly (monthly or more) | 12 | 44.4 |
| **How far is the nearest HIV prevention service center from your location** |  |  |
| 1-5 km | 11 | 40.7 |
| Less than 1 km | 9 | 33.4 |
| More than 5 km | 7 | 25.9 |
| **Challenges faced when accessing HIV prevention services (Select all that apply)** |  |  |
| Distance to service centers | 7 | 17.1 |
| Stigma or discrimination | 10 | 24.4 |
| Cost of services | 9 | 21.9 |
| Lack of information about services | 15 | 36.6 |

**Table 4:**
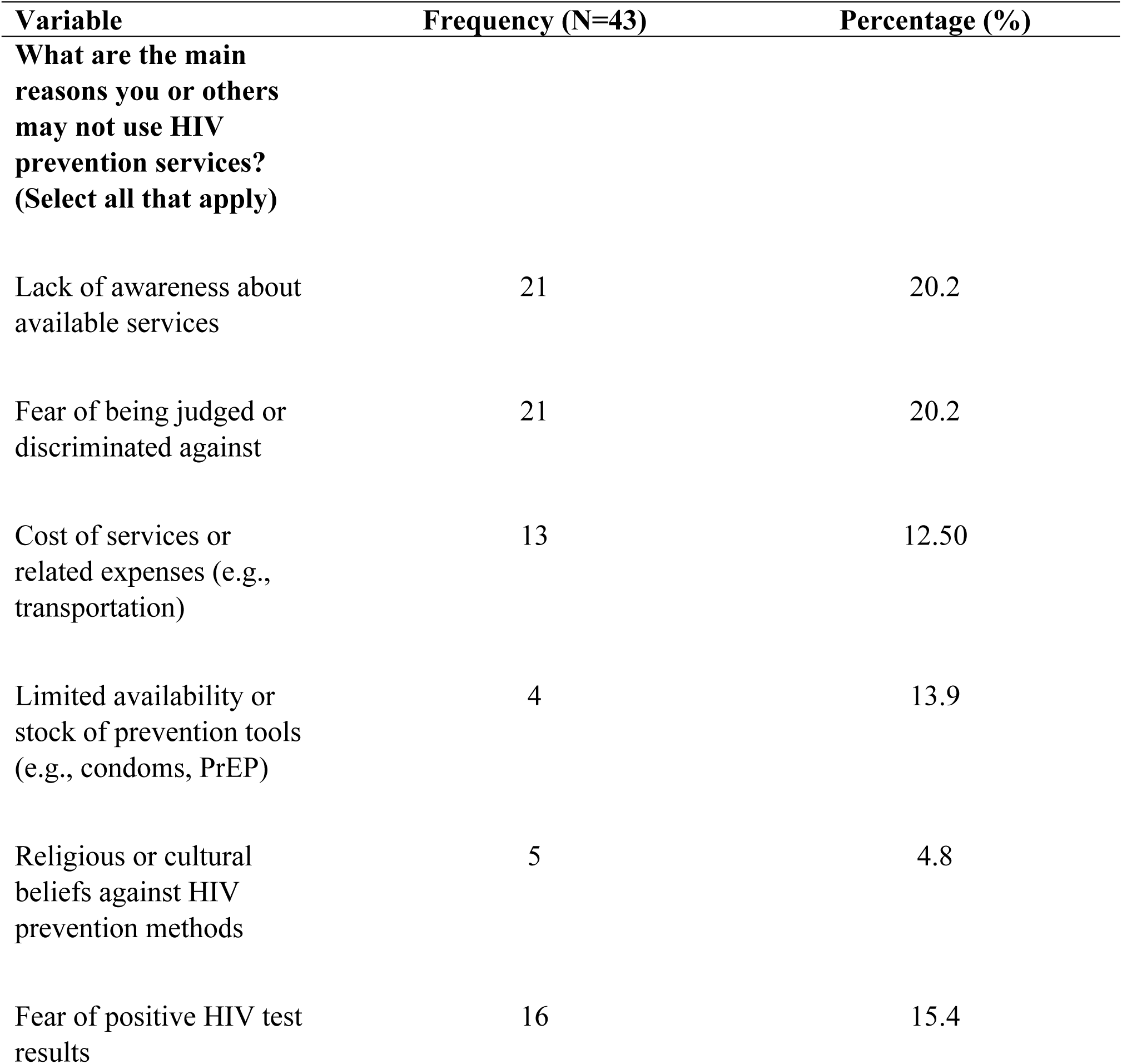
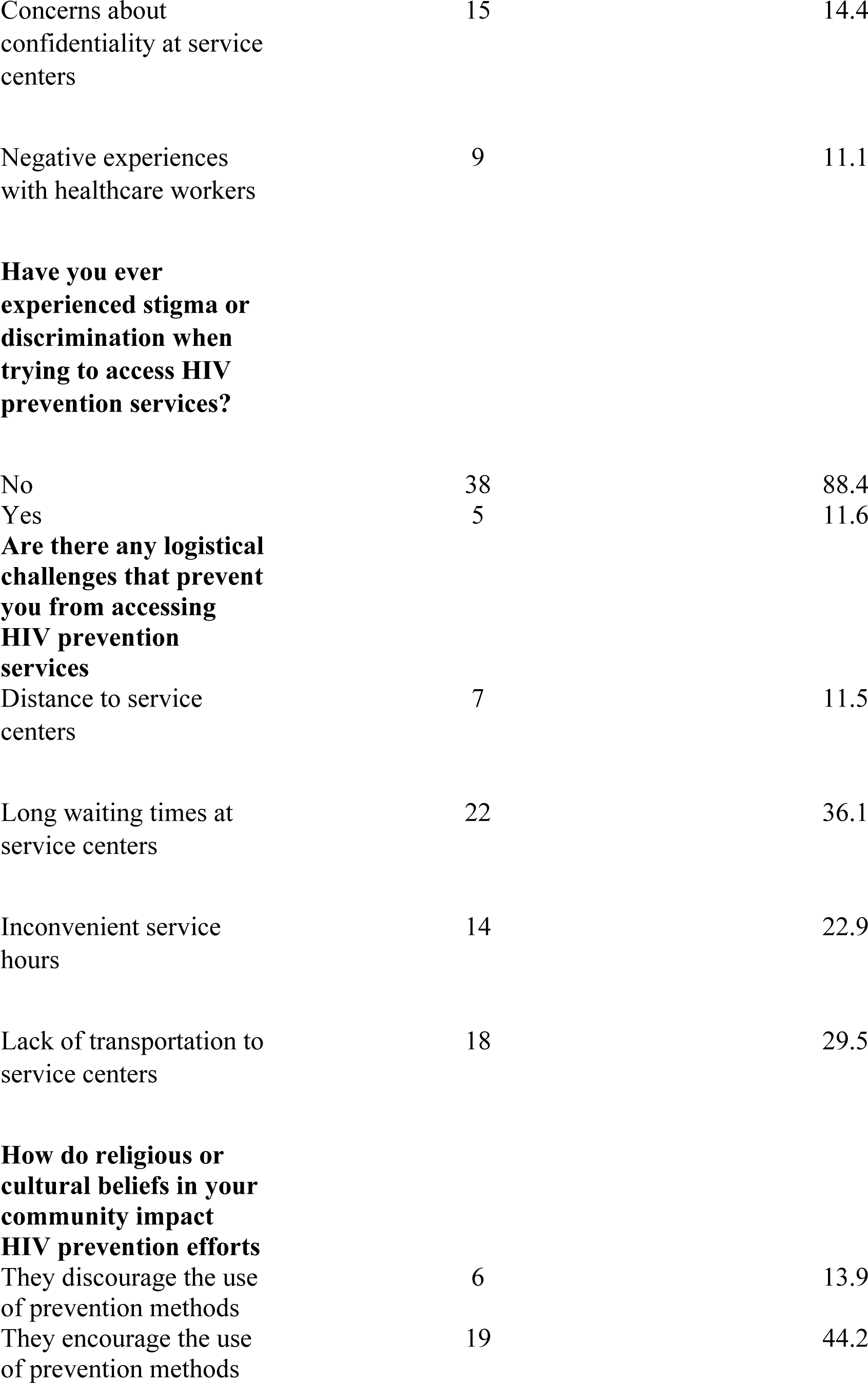
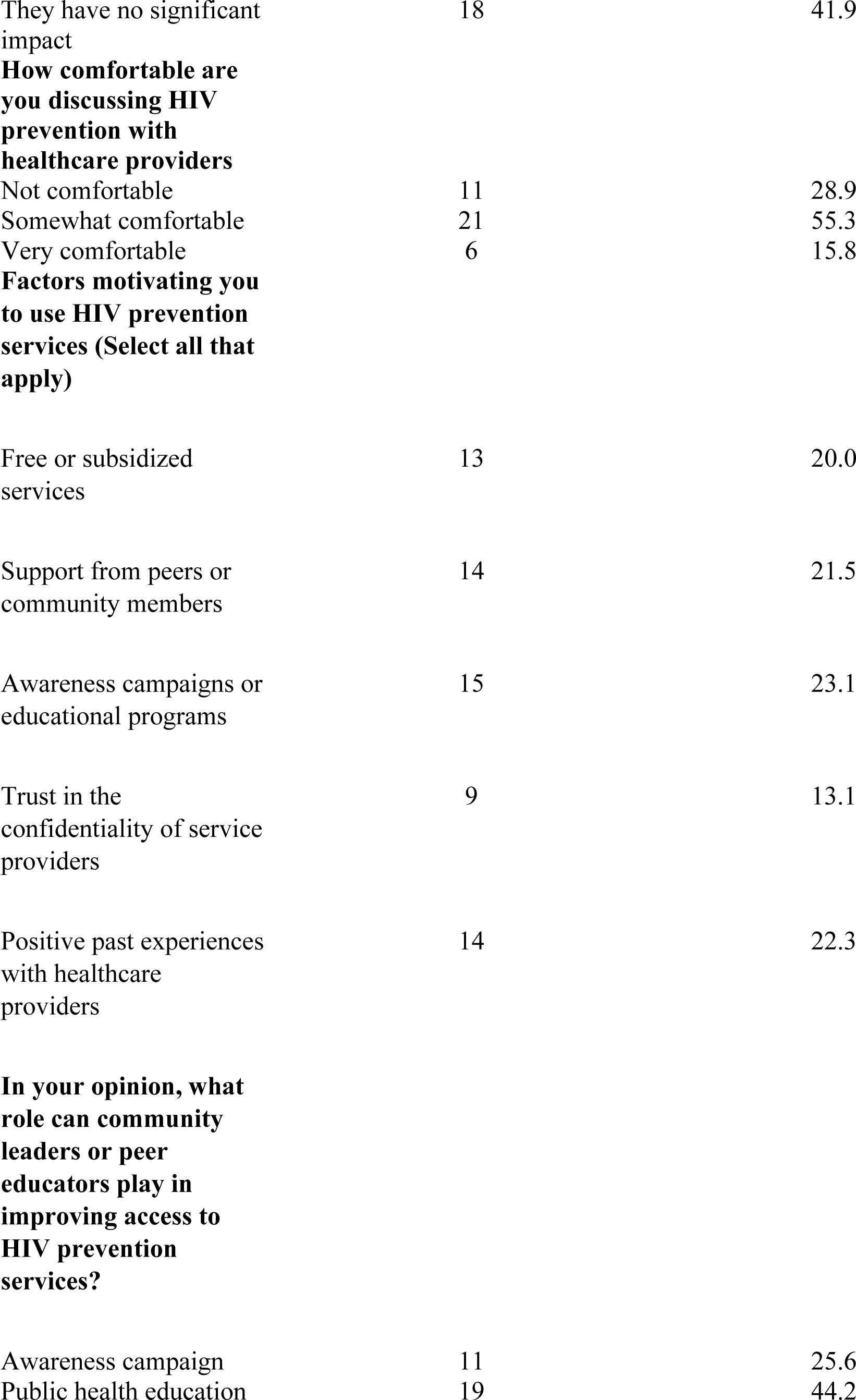
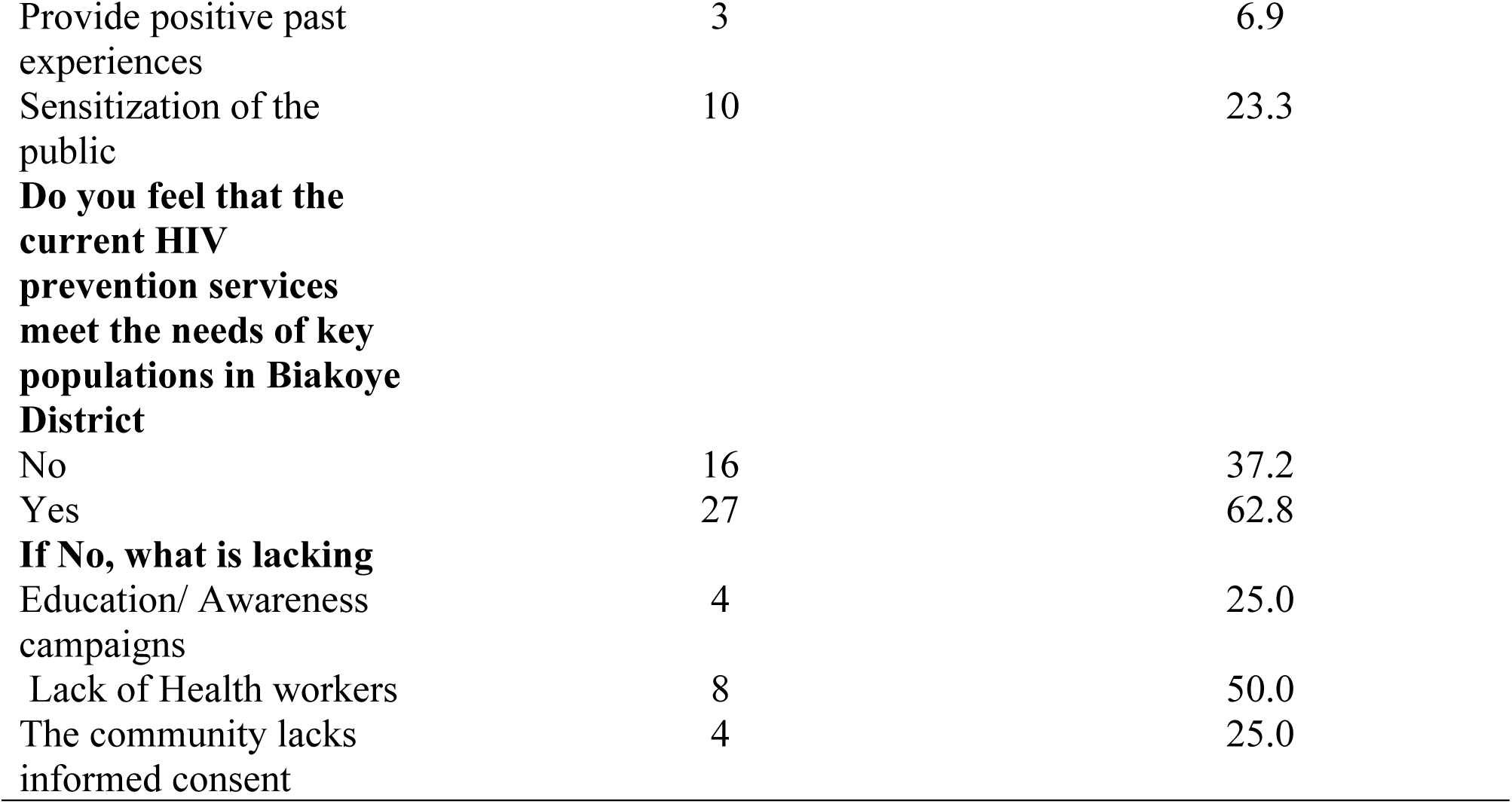
Barriers and Facilitators to Prevention.

| Variable | Frequency (N=43) | Percentage (%) |
| --- | --- | --- |
| **What are the main reasons you or others may not use HIV prevention services? (Select all that apply)** |  |  |
| Lack of awareness about available services | 21 | 20.2 |
| Fear of being judged or discriminated against | 21 | 20.2 |
| Cost of services or related expenses (e.g., transportation) | 13 | 12.50 |
| Limited availability or stock of prevention tools (e.g., condoms, PrEP) | 4 | 13.9 |
| Religious or cultural beliefs against HIV prevention methods | 5 | 4.8 |
| Fear of positive HIV test results | 16 | 15.4 |
| Concerns about confidentiality at service centers | 15 | 14.4 |
| Negative experiences with healthcare workers | 9 | 11.1 |
| **Have you ever experienced stigma or discrimination when trying to access HIV prevention services?** |  |  |
| No | 38 | 88.4 |
| Yes | 5 | 11.6 |
| **Are there any logistical challenges that prevent you from accessing HIV prevention services** |  |  |
| Distance to service centers | 7 | 11.5 |
| Long waiting times at service centers | 22 | 36.1 |
| Inconvenient service hours | 14 | 22.9 |
| Lack of transportation to service centers | 18 | 29.5 |
| **How do religious or cultural beliefs in your community impact HIV prevention efforts** |  |  |
| They discourage the use of prevention methods | 6 | 13.9 |
| They encourage the use of prevention methods | 19 | 44.2 |
| They have no significant impact | 18 | 41.9 |
| **How comfortable are you discussing HIV prevention with healthcare providers** |  |  |
| Not comfortable | 11 | 28.9 |
| Somewhat comfortable | 21 | 55.3 |
| Very comfortable | 6 | 15.8 |
| **Factors motivating you to use HIV prevention services (Select all that apply)** |  |  |
| Free or subsidized services | 13 | 20.0 |
| Support from peers or community members | 14 | 21.5 |
| Awareness campaigns or educational programs | 15 | 23.1 |
| Trust in the confidentiality of service providers | 9 | 13.1 |
| Positive past experiences with healthcare providers | 14 | 22.3 |
| **In your opinion, what role can community leaders or peer educators play in improving access to HIV prevention services?** |  |  |
| Awareness campaign | 11 | 25.6 |
| Public health education | 19 | 44.2 |
| Provide positive past experiences | 3 | 6.9 |
| Sensitization of the public | 10 | 23.3 |
| **Do you feel that the current HIV prevention services meet the needs of key populations in Biakoye District** |  |  |
| No | 16 | 37.2 |
| Yes | 27 | 62.8 |
| **If No, what is lacking** |  |  |
| Education/ Awareness campaigns | 4 | 25.0 |
| Lack of Health workers | 8 | 50.0 |
| The community lacks informed consent | 4 | 25.0 |

**Table 5:** Factors associated with accessibility and utilization of HIV services and marginalized population.

| Variables | Accessibility and Utilization of HIV Services |  | cOR ((95% CI), p-value | aOR(95% CI), p-value |
| --- | --- | --- | --- | --- |
|  | Low (16)n (%) | High (27)n (%) |  |  |
| Level of education |  |  |  |  |
| No formal education | 5(31.3) | 1(3.7) | Ref. | Ref. |
| Primary education | 4(25.0) | 4(14.8) | 1.30(3.53-9.29), 0.253 | 1.30(3.53-9.29), 0.253 |
| Secondary education | 4(25.0) | 10 (37.0) | 2.15 (0.03-4.26), 0.047 | 2.15 (0.03-4.26), 0.047 |
| Tertiary education | 3(18.7) | 12(44.5) | 2.60 (0.41-4.73), 0.020 | 2.60 (0.41-4.73), 0.020 |
| No | 10(62.5) | 5(18.5) | Ref. | Ref. |
| Yes | 6(37.5) | 22(81.5) | 1.89 (0.54-3.24), 0.006 | 1.55(0.08-3.03), 0.039 |
| How confident are you in your knowledge of HIV prevention methods |  |  |  |  |
| Not confident | 5(31.3) | 4(14.8) | Ref. | Ref. |
| Somewhat confident | 11(68.7) | 13(48.2) | 0.36 (1.11-1.83), 0.631 | 0.14 (1.51-1.78), 0.872 |
| Very confident | 0(0.0) | 10(37.0) | 3.25 (0.15-6.34), 0.040 | 2.51(0.68-5.71), 0.123 |

### Knowledge of HIV Prevention

Overall, 93.0% had heard of HIV prevention methods. Condoms (68.5%) and regular testing (22.2%) were the most recognized methods, while only 9.3% were aware of PrEP. Confidence in knowledge varied: 23.3% were very confident, 55.8% somewhat confident, and 20.9% not confident.

### Accessibility and Utilization of HIV Prevention Services

A total of 62.8% had ever used HIV prevention services. Condoms (71.1%) and HIV testing (26.3%) were the most used services. Only one participant (2.6%) reported using PrEP.

Accessibility varied: 33.4% lived <1 km from a service center, 40.7% lived 1–5 km away, and 25.9% lived >5 km away.

### Barriers and Facilitators to HIV Prevention

Participants identified multiple barriers to accessing HIV prevention services. The most commonly reported barriers included lack of awareness of available services (48.8%), fear of stigma or discrimination (48.8%), long waiting times at service centers (36.1%), lack of transportation (29.5%), fear of receiving a positive HIV test result (37.2%), and concerns about confidentiality (34.9%). Cost of services was also reported by 21.9% of participants. Facilitators to service utilization included community awareness campaigns (34.9%), support from peers or community members (32.6%), positive past experiences with healthcare providers (32.6%), availability of free or subsidized services (30.2%), and trust in the confidentiality of service providers (20.9%).

### Factors Associated with Utilization of HIV Prevention Services

In the multivariable logistic regression analysis, two factors were significantly associated with the utilization of HIV prevention services. Participants with tertiary education had higher odds of using HIV prevention services compared to those with no formal education (aOR = 2.60, 95% CI: 0.41–4.73, p = 0.020). Similarly, participants who knew where to access HIV prevention services were more likely to utilize them than those who did not (aOR = 1.55, 95% CI: 0.08–3.03, p = 0.039). No significant associations were observed with age, sex, marital status, or comfort discussing HIV prevention with healthcare providers

## DISCUSSION

This study explored barriers and facilitators to HIV prevention among key populations in a rural Ghanaian district. Awareness of HIV prevention was high, aligning with previous studies in sub-Saharan Africa (10,11). However, awareness of PrEP was notably low, consistent with studies showing limited PrEP rollout and education among KPs in Ghana and similar settings (12,13).

Despite high awareness, actual utilization of prevention services was modest. Only 62.8% had ever used HIV prevention services, and PrEP use was nearly absent. This gap between knowledge and consistent preventive behavior is widely documented in KP research (14,15).

Barriers identified stigma, discrimination, confidentiality concerns, long waiting times, transportation challenges, and inadequate awareness mirror findings from other African contexts (16–19). Stigma remains one of the strongest deterrents to service uptake.

Higher education and knowledge of service locations were significant predictors of service utilization. Education enhances health literacy and the ability to navigate health systems, consistent with previous research (20). Awareness of service locations improves accessibility and reduces uncertainty, supporting earlier evidence (21–23).

Facilitators such as peer support, awareness campaigns, free services, and positive provider experiences underscore the value of KP-friendly and community-driven approaches. Evidence consistently shows that peer-led programs and community empowerment strengthen prevention engagement (16,23,24).

These findings highlight the need for flexible, community-based HIV prevention models in rural areas like Biakoye, where structural barriers impede access.

## CONCLUSION

Although awareness of HIV prevention is high among key populations in Biakoye District, consistent utilization remains limited due to social and structural barriers. Education level and knowledge of service locations significantly influence service uptake. Strengthening community-based strategies, reducing stigma, improving service accessibility, and expanding PrEP education are essential for improving HIV prevention outcomes.

## Data Availability

All relevant data are within the manuscript and its Supporting Information files.

## ACKNOWLEDGMENTS

The authors thank all study participants and the Biakoye District Health Directorate for their support. Appreciation is extended to the data collectors and field assistants who contributed to recruitment and questionnaire administration.

## DECLARATIONS

### Ethics approval and consent to participate

Approved by the Ghana Health Service Ethics Review Committee (GHS-ERC: 022/03/25). Written informed consent obtained.

### Consent for publication

Not applicable.

### Availability of data and materials

Available from the corresponding author upon reasonable request.

### Authors’ contributions

EKH: Conceptualization, data collection, analysis, manuscript drafting

RAW: Data collection, field coordination

MT: Data analysis, interpretation

MPK: Supervision, guidance, manuscript review

All authors approved the final manuscript.

